# Genotypic analysis of *Plasmodium falciparum* malaria parasites in a clinical trial of RTS,S vaccination in combination with seasonal malaria chemoprevention

**DOI:** 10.1101/2025.11.21.25340773

**Authors:** Emily LaVerriere, Katrina Kelley, Alassane Dicko, Issaka Sagara, Almahamoudou Mahamar, Jean-Bosco Ouedraogo, Issaka Zongo, Halidou Tinto, Daniel Chandramohan, Brian Greenwood, Daniel E. Neafsey

## Abstract

Both vaccination and seasonal chemoprevention are effective interventions against malaria, but both morbidity and mortality due to malaria remain high, particularly in sub-Saharan Africa. A recent clinical trial showed that the combination of these interventions provided substantial protection against malaria over a five-year period. We performed amplicon-based genotyping of highly polymorphic parasite antigens within 1,530 samples from this clinical trial. We evaluated the complexity of infection within these samples, finding that participants who received both interventions had less complex infections (fewer genetically distinct parasite strains) than participants who received either intervention given alone. We also evaluated the prevalence of parasites matching the vaccine construct CSP, at the level of haplotypes, epitopes, and individual amino acid changes. We found no significant differences in the prevalence of vaccine-matching alleles between vaccinated and unvaccinated groups. For context, this study genotyped about half as many non-vaccinated participants as the original phase 3 trial for RTS,S. While a comparable number of vaccinated individuals were genotyped in both studies, the infections in this study had less complex infections, leading to an overall lower number of parasite strains to analyze. These results suggest that the combination of therapies provides protection from infection, in addition to clinical disease, and they highlight the importance of continued molecular surveillance as malaria interventions are deployed.

## Introduction

Malaria remains a persistent global health threat, causing an estimated 597,000 deaths per year, primarily in children under five years of age (1). The burden of malaria is greatest in sub-Saharan Africa, where in many regions disease transmission is seasonal in nature due to rainfall-associated changes in the population size of *Anopheles spp.* mosquitoes that transmit the disease (2). While immunity to clinical disease has been demonstrated to accrue with age, acquisition of asymptomatic infections is age-independent, and occurs with similar timing across age groups following the commencement of the rainy season (3,4).

Effective interventions for malaria in seasonal transmission settings are therefore timed to coincide with increased transmission, and include seasonal chemoprevention (SMC) and, more recently, seasonal vaccination. Seasonal chemoprevention usually involves multiple, regularly administered doses of sulfadioxine-pyrmethamine and amodiaquine during the expected transmission season (5). Multiple vaccine candidates and schedules have been tested (6,7), and the RTS,S/AS01_E_ and R21 vaccines are currently recommended for use by the WHO in malaria-endemic areas, each with a recommended three-dose primary schedule, followed by a booster dose (8,9). The timing of administration of the booster dose is usually age based but countries with highly seasonal malaria transmission may consider giving the booster dose prior to the malaria transmission season (9). Both SMC and vaccination provide protection against disease that wanes over time. A recent clinical trial compared the combination of SMC and seasonal vaccination with the RTS,S/AS01_E_ vaccine (subsequently termed RTS,S) to each intervention on its own in areas with strongly seasonal malaria transmission (10,11). In this trial, the combination of SMC with RTS,S significantly reduced the incidence of uncomplicated disease, severe disease, and death compared to either intervention on its own.

In this study, we performed genotyping of highly polymorphic parasite antigens in participant samples obtained from children in each of the three arms of the combined SMC and vaccination clinical trial (RTS,S alone, SMC alone, and the combination) in order to understand the nature of protection of the individual and combined interventions against infection. Our objectives were to determine whether the vaccine provided differential protection against parasites that had an exact match at the circumsporozoite protein (CSP) antigen to the RTS,S vaccine construct, and to estimate the number of genomically distinct parasite lineages within each sample as a measure of protection against infection. Previous large-scale genotyping studies have examined the parasites present in infections from multiple RTS,S clinical trials (12,13). The initial genotyping analysis of the phase 3 trial found increased vaccine efficacy against parasite strains matching the CSP genotype used within the vaccine. More recent work analyzing a phase 2b clinical trial comparing different vaccine dosing regimens found that all vaccine regimens tested blocked some parasite infections, and that complexity of infection was reduced in participants in the RTS,S arm compared to the control arm.

In this study, we generated parasite genotyping data from samples meeting the clinical case definition for malaria from all three arms of the combined SMC and vaccination clinical trial (10,11), allowing us to profile the number of clones in each. While the original study demonstrated a clear effect of the combined interventions on clinical disease, these newly-generated genotyping data demonstrate evidence of combined efficacy against infection in a non-strain-specific manner.

## Methods

### Samples / original trial

We processed samples from the clinical trial NCT04319380, registered on ClinicalTrials.gov, as previously described (10,11). This study was a double-blind, individually-randomized, controlled, non-inferiority and superiority phase 3 trial done at two sites: the Bougouni district and neighboring areas in Mali and Houndé district, Burkina Faso. The original trial protocol was reviewed and approved by the ethics committees of the London School of Hygiene & Tropical Medicine, UK; the University of Science, Techniques, and Technologies, Bamako, Mali; the ethics committee for health research, Burkina Faso; and the regulatory authorities in Burkina Faso and Mali.

Briefly, 6,861 children 5 to 17 months of age were enrolled and randomly assigned to receive either sulfadoxine-pyrimethamine and amodiaquine (the chemoprevention-alone group, 2,287 children), RTS,S/AS01_E_ (the vaccine-alone group, 2,288 children), or both therapies (the combination group, 2,286 children). The chemoprevention-alone group received tetanus or tetanus-diphtheria toxoid vaccine in place of RTS,S/AS01_E_, and the vaccine-alone group also received sulfadoxine-pyrimethamine and amodiaquine placebos. Participants each received five doses of RTS,S/AS01_E_ or placebo vaccination: three in April-June 2017, followed by annual doses in 2018 and 2019. Participants each received four courses of sulfadoxine-pyrimethamine and amodiaquine or placebo per year. All participants received a long-lasting insecticide-treated bed net when enrolled into the study.

Uncomplicated malaria was the primary outcome of the original trial, defined as fever (either measured at least 37.5LJ or fever within the previous 48 hours) and *P. falciparum* parasitemia of at least 5,000 parasites per mm^3^. Participants presenting to trial health facilities with suspected malaria were tested with rapid diagnostic tests, and those with positive tests received artemether-lumefantrine and gave a blood sample for microscopic confirmation.

The initial trial lasted three years (10), and participants who did not have severe reactions to the interventions (febrile convulsions after vaccination) were invited to re-enroll in the extension study for the subsequent two years (11). In 2020, 5098 children consented to re-enroll in the extension study; these participants retained the same treatment groups as before, resulting in 1,683, 1,705, and 1,710 participants in the extension study combination group, vaccine-only group, and chemoprevention-only groups, respectively. Participants no longer received SMC when they reached the age of five years, in line with national guidelines, and seasonal vaccination was no longer given after they reached this age. Thus, participants who enrolled when aged 5-11 months received two additional doses of RTS,S/AS01_E_ or placebo vaccination in June 2020 and June 2021, as well as four cycles of sulfadoxine-pyrimethamine and amodiaquine or placebo per year in 2020 and 2021, whilst those enrolled when aged 12-17 months received only one additional year of SMC and seasonal vaccination in 2020.

A 1:2 random sample from all samples which came from infections meeting the clinical definition of malaria in the original trial (symptomatic disease with ≥5,000 parasites/μL identified via microscopy) was selected for genotyping across intervention and country for each year (2017, 2019, 2021). This selection resulted in 730 samples from the vaccine-only group, 572 samples from the chemoprevention-only group, and 228 samples from the combination group (**Table 1**). Sequencing was attempted on all selected samples.

**Table 1.**

| Country | Burkina Faso |  |  | Mali |  |  |
| --- | --- | --- | --- | --- | --- | --- |
| Year | RTS,S | SMC | RTS,S+SMC | RTS,S | SMC | RTS,S+SMC |
| 2017 | 114 | 50 | 12 | 85 | 76 | 10 |
| 2019 | 204 | 182 | 61 | 65 | 82 | 37 |
| 2021 | 154 | 71 | 58 | 108 | 111 | 50 |

### Sequencing

We extracted genomic DNA from the dried blood spot samples and prepared them for sequencing using the 4CAST amplicon panel, as previously described (14), with no changes in protocol from the original publication. This amplicon panel targets four highly polymorphic regions of the *P. falciparum* genome, within the *csp, ama1, sera2*, and *trap* genes. Particularly relevant for this study, the amplicon within *csp* covers the polymorphic C-terminal region that is part of the RTS,S vaccine construct. The coverage of this region of *csp* allows for direct identification of parasites matching or mis-matching the vaccine strain in this region. The high diversity captured within this amplicon panel also allows estimation of complexity of infection (COI; number of genotypically distinct strains within an infection) more accurately than a single amplicon alone.

We sequenced the majority of samples (n=1,542) on two lanes of Illumina NovaSeq instruments, with 768 samples per lane. We sequenced a second batch of samples (n=192) on an Illumina MiSeq instrument. All sequencing runs were paired-end, 2×250 bp reads. Samples were processed in 96-well plates, with each plate containing extraction and PCR controls that were carried through sequencing and analysis. Data from these samples were submitted to the NCBI Sequencing Read Archive (http://www.ncbi.nih.gov/sra) under accession PRJNA1345094.

### Analysis

We processed the data through a previously described pipeline (14), designed for analyzing deeply sequenced multiplexed amplicon sequencing data and parsing it into haplotypes. We set a threshold for the minimum number of reads per haplotype per sample at 200 reads (for samples sequenced on NovaSeq) or 20 reads (for samples sequenced on MiSeq). We also required a haplotype to have a within-sample frequency of at least 1% as estimated by sequencing reads, and we removed any singleton haplotypes from the full dataset. We examined the positive and negative control wells present on every 96-well plate; control wells were required to have the included strain (Dd2) detected, and negative controls were required to have no haplotypes with reads above 200 (for NovaSeq samples) or 20 (for MiSeq samples). The negative control in one 96-well plate did not pass these checks, so all samples on that plate were removed from the final dataset.

One of the two NovaSeq lanes had several sequencing artifacts. Specifically, three nucleotide changes within the CSP amplicon were detected in some samples (genomic coordinates 221383, 221403, and 221412, all changed to C). We inspected this region in previously generated amplicon sequencing datasets of parasites from similar regions, and none of these three changes appeared (3,15). We also looked within the Pf3k dataset of whole-genome data (16) and did not see these nucleotide changes in any samples. Finally, we selected eight samples where these nucleotide changes were present, re-amplified this region with the CSP primers, and confirmed with Sanger sequencing that the nucleotide changes were not present in the samples. We masked these three positions in all downstream analysis.

We performed all subsequent analyses on denoised haplotype calls in R 4.2.0, with the packages Biostrings, DescTools, grantham, here, RColorBrewer, stats, and tidyverse (17–23). We estimated COI as the maximum number of distinct haplotypes identified at an individual 4CAST locus (CSP, AMA1, SERA2, TRAP) for an individual sample. All statistical tests were two-sided, unless otherwise noted. We tested for significance using the “kruskal.test”, “fisher.test”, and “p.adjust” functions from the stats package, as well as the “BinomDiffCI” function from the DescTools package.

## Results

### Infections are less complex in individuals who received RTS,S/AS01_E_ with SMC

We generated high-quality *P. falciparum* sequencing data from 1,530 samples in total, all of which came from infections meeting the clinical definition of malaria in the original trial: symptomatic disease with ≥5000 parasites/μL identified via microscopy. Concordant with the original study (10), the number of clinical disease cases was much lower in the individuals who received both SMC and RTS,S, leading to an uneven number of samples to sequence between each study arm. Thus, we sequenced 228 samples from the RTS,S with SMC arm, compared to 730 samples from the RTS,S arm and 572 samples from the SMC arm (**Table 1**).

We first leveraged the genotyping data to identify complex (polyclonal) infections. We estimated the complexity of infection, or the number of genetically distinct parasites identified in each sample. We found that monoclonal infections were more common in individuals who received both RTS,S and SMC than in those who received SMC only (**Figure 1A**; Kruskal-Wallis test, x^2^(2) = 9.11, P = 0.011; Dunn test with Holm correction, both vs. SMC only: adjusted P = 0.0079).

**Figure 1.**
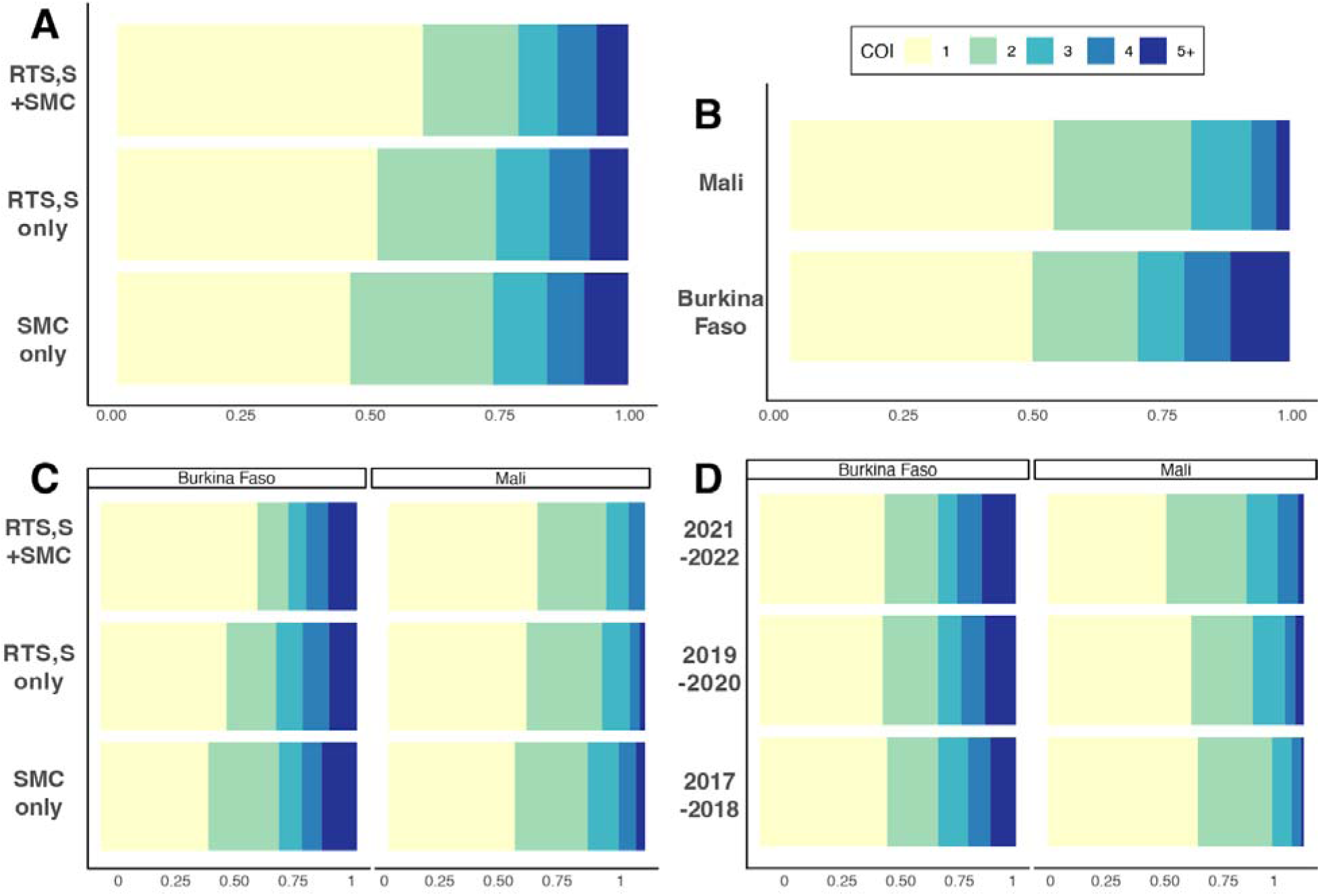
All panels visualize the proportion of samples at estimated complexity of infection (COI) level, from 1 (monoclonal) to 5+ (highly polyclonal). A) Samples are stratified by study arm. Samples are more likely to have lower COI in the combination-therapy group (Kruskal-Wallis test, x^2^(2) = 9.11, P = 0.011; Dunn test with Holm correction, both vs. SMC only: adjusted P = 0.0079). B) Samples stratified by country. COI tended to be higher in samples in Burkina Faso than in samples from Mali (Kruskal-Wallis test, x^2^(1) = 22.35, P = 1.41 e^-5^; mean COI in Burkina Faso = 2.3, mean COI in Mali = 1.8). C) Samples stratified by study arm and country. Samples are significantly less complex in the combination group in Burkina Faso (Kruskal-Wallis test, x^2^(2) = 6.84, P = 0.03; Dunn test with Holm correction, both vs. SMC only: adjusted P = 0.027). Samples are less complex in the combination group in Mali, though not significantly (Kruskal-Wallis test, x^2^(2) = 3.76, P = 0.15). D) Samples stratified by time period and country. COI distributions were consistent over time in Burkina Faso (Kruskal-Wallis test, x^2^(2) = 0.22, P = 0.90), but differed in later years in Mali (Kruskal-Wallis test, x^2^(2) = 8.85, P = 0.011; Dunn test with Holm correction, 2017-2018 vs. 2021-2022, adjusted P = 0.012).

We also compared COI distributions by country (**Figure 1B**). Both countries had similar proportions of monoclonal infections - approximately half of all sequenced infections were monoclonal (48.5% in Burkina Faso (n=393), 52.3% in Mali (n=390). However, among the infections with more than one clone detected, COI tended to be higher in individuals in Burkina Faso than in individuals from Mali (Kruskal-Wallis test, x^2^(1) = 22.35, P = 1.41 e^-5^; mean COI in Burkina Faso = 2.3, mean COI in Mali = 1.8). When stratified by both study arm and country (**Figure 1C**), COI distributions follow a similar trend as in **Figure 1A**. In Burkina Faso, the pattern of monoclonal infections being more common in the RTS,S + SMC study arm was observed again (Kruskal-Wallis test, x^2^(2) = 6.84, P = 0.03; Dunn test with Holm correction, both vs. SMC only: adjusted P = 0.027). Monoclonal infections remained more common in RTS,S + SMC individuals in Mali as well, though the trend did not reach the threshold of statistical significance (Kruskal-Wallis test, x^2^(2) = 3.76, P = 0.15).

COI distributions remained consistent over time in Burkina Faso (**Figure 1D**; Kruskal-Wallis test, x^2^(2) = 0.22, P = 0.90). In Mali, monoclonal infections became less frequent in later years (Kruskal-Wallis test, x^2^(2) = 8.85, P = 0.011; Dunn test with Holm correction, 2017-2018 vs. 2021-2022, adjusted P = 0.012).

### Parasites matching the vaccine strain appear at similar proportions in all groups

We compared the prevalence of CSP haplotypes matching the 3D7 vaccine strain between participants who received RTS,S and those who did not (**Figure 2a**). Prevalence of amino acid haplotypes matching 3D7 did not differ between vaccinated participants (127 3D7-matching infections, prevalence = 14.4%) and unvaccinated participants (81 3D7-matching infections, prevalence = 15.1%; Fisher’s exact test, one-sided, P = 0.386). Additionally, we found no difference when considering the frequency of 3D7-matching haplotypes in place of prevalence; 3D7-matching haplotypes in vaccinated participants had a prevalence of 8.73% among all haplotypes, compared to a prevalence of 8.56% among non-vaccinated participants (Fisher’s exact test, one-sided, P = 0.579). Full lists of nucleotide and amino acid haplotypes observed in participants stratified by vaccination status are shown in **Supplemental Tables 1-2**. We also considered how similar the common haplotypes were to 3D7, by summing the Grantham distance of each mismatched amino-acid in a given haplotype (**Figure 2b**). None of the most common haplotypes were very similar to 3D7 (sum of Grantham distances ranged from 319 to 533 per haplotype).

**Figure 2.**
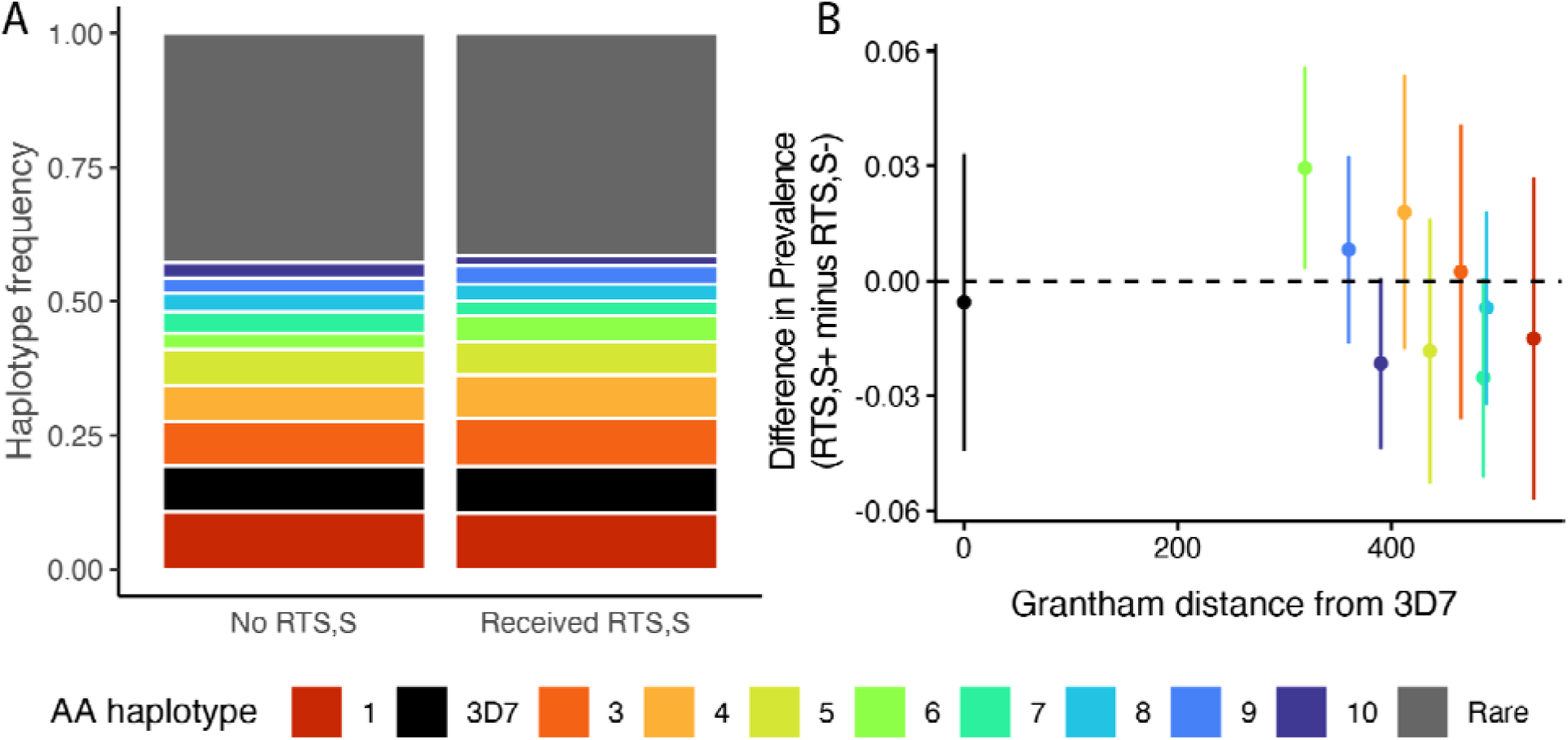
A) Frequency of the top ten most common amino acid CSP haplotypes (each with a distinct color), including the 3D7-matching haplotype (second most common, colored in black). All less common haplotypes are grouped together and colored in grey. 3D7-matching haplotype prevalence did not differ between vaccinated and unvaccinated participants (Fisher’s exact test, one-sided, P = 0.386). B) Grantham distance for each haplotype compared to the 3D7 haplotype is on the x-axis (distance is a sum of the Grantham distance from each non-3D7-matching amino acid per haplotype). The difference in prevalence of the haplotype between vaccinated and unvaccinated groups is on the y-axis. The lines spanning each point plot the 95% confidence interval around the difference in prevalences.

We also evaluated whether evidence of allele-specific protection could be identified at the three epitopes previously described for CSP: DV10, Th2R, and Th3R, rather than at the full haplotype level (**Table 2**). We compared the prevalence of amino acid haplotypes spanning each of these epitope regions individually, and we again found no difference associated with vaccination status forTh2R, Th3R, or DV10 (Fisher’s exact tests, one-sided; P = 0.641, 0.609, 0.778, respectively). The incidence of each epitope haplotype per study arm is described in

**Table 2.**

| Locus | 3D7-matching allele prevalence |  |  | P-value |
| --- | --- | --- | --- | --- |
|  | RTSS+ | RTSS- | Difference |  |
| CSP | 0.147 | 0.152 | -0.005 | 0.386 |
| Th2R | 0.157 | 0.162 | -0.005 | 0.641 |
| Th3R | 0.214 | 0.223 | -0.009 | 0.609 |
| DV10 | 0.177 | 0.173 | 0.004 | 0.778 |

### Supplemental Tables 3-5

Finally, we investigated individual amino acid positions within these epitopes to look for evidence of 3D7-specific protection. Specifically, we examined amino acid positions 299, 301, 317, 354, 356, 359, 361, which all vary in these populations, consistent with previous deep sequencing studies of CSP (13,15). We examined each position on its own, and we compared the distributions of amino acids observed at that position across each of the study arms. The prevalence of the 3D7-matching allele at position 321 was higher in vaccinated participants (n = 281, prevalence = 0.319) compared to unvaccinated participants (n = 151, prevalence = 0.283. While the initial hypothesis testing was significant (Fisher’s exact test, one-sided, P = 0.01), after adjusting for multiple hypothesis testing, the result is on the edge of significance (adjusted P = 0.22, after Benjamini-Hochberg correction). None of the other distributions varied significantly from each other, even before correcting for multiple hypothesis testing (Fisher’s exact test applied to each residue, prevalences and p-values in **Supplemental Table 6**, raw number of appearances of each residue in **Supplemental Table 7**).

## Discussion

In this study, we generated amplicon sequencing data to explore parasite diversity within symptomatic infections in a clinical trial which compared the combination of SMC with seasonal vaccination with RTS,S with either intervention given alone (10). Genotyping data has the capacity to both confirm and, in this case, recapitulate the efficacy of the interventions in the trial. While the original study found an increase in protection against symptomatic disease in those who received both SMC and vaccination (10,11), we found an additional benefit of the combined interventions. Participants who received both interventions were also more likely to exhibit lower complexity infections than those receiving only SMC or RTS,S alone. This suggests that participants who received both interventions were more effective at blocking a subset of infections, whether polyclonal infections were founded via multiple infectious mosquito bites or single polyclonal infectious bites. This finding suggests that the combined chemoprevention and vaccination not only protects against clinical disease, but also has additive protection against parasite infection itself, which can occur regardless of clinical symptoms. This protection may manifest as lower complexity infections, as previously seen in individuals vaccinated with RTS,S (12).

The genotyping data generated also allowed us to look for differential protection as a result of vaccination, which was observed in the original phase 3 trial of the RTS,S/AS01_E_ vaccine (13). We did not observe vaccination-related differences in the prevalence of 3D7-matching alleles when considering full haplotypes or epitopes. Two factors may have resulted in a greater temporal lag and a smaller impact of vaccination on the observed breakthrough infections genotyped here compared to the phase 3 trial. First, the administration of SMC in the combination group may have delayed the time to first infection within each season of transmission, thus leading to first infections occurring when vaccine efficacy has started to decrease. Second, the samples genotyped in this study were a random selection of all symptomatic cases (stratified by country and year), compared to the phase 3 trial which genotyped the first post-vaccination infection per individual.

We did observe increased prevalence of the 3D7-matching CSP amino acid 321N in vaccinated individuals, although this difference did not reach statistical significance after correcting for multiple hypothesis testing. However, we cannot rule out that a smaller magnitude difference in protection could be present below our limit of detection. With a minimum of 500 samples per group, estimated prevalence of the 3D7-matching haplotype of 0.15 in the unvaccinated population, and a significance threshold of 0.05, this study had only 80% power to detect a difference of ∼0.05 in prevalence between groups. Any smaller differences in prevalence would require higher sample size. As vaccination rates increase throughout malaria-endemic regions, continued surveillance of malaria infections in vaccinated and unvaccinated populations will be necessary for detecting any signals of vaccine escape, and larger studies will be better powered to estimate the magnitude of any allele-specific protective efficacy that is present.

## Conclusion

Combining seasonal malaria chemoprevention with vaccination protected against clinical disease in a previous clinical trial. In the study reported in this paper, we found that the combination of SMC and seasonal vaccination also led to reduced levels of polyclonality within samples from participants with clinical disease, indicating increased protection against infection itself relative to participants receiving only one intervention. Infection polyclonality was similar between the RTS,S-only and SMC-only study, suggesting comparable protection against infection provided by each intervention given alone, as was noted in analysis of clinical infections. These genotyping results underscore the additive value of vaccination and SMC for protecting children against malaria in seasonal transmission settings. In the RTS,S-only and RTS,S+SMC study arms we found no evidence of significantly greater protection against parasites with CSP C-terminus sequences matching the 3D7 vaccine strain, suggesting that such protection is modest in magnitude if present. Nevertheless, we advocate continued surveillance for evidence of vaccine escape as RTS,S and R21 are more widely deployed in Africa, when power to detect allele-specific protection may be enhanced.

## Data Availability

All genotyping data produced has been submitted to the NCBI Sequencing Read Archive (http://www.ncbi.nih.gov/sra) under accession PRJNA1345094.

## Acknowledgements

We thank Sanni Ali and Paul Snell for their assistance with sample selection and coordination.

## Funding Declaration

The overall trial on which the study described in this paper and the laboratory analyses described was funded by the UK Joint Global Health Trials (Department of Health and Social Care (DHSC), the Foreign, Commonwealth & Development Office (FCDO), the Global Challenges Research Fund (GCRF), the Medical Research Council (MRC) and the Wellcome Trust (WT)) (grant MR/V005642/1). This UK funded award was part of the EDCTP2 programme supported by the European Union. The trial was also funded by PATH’s Malaria Vaccine Initiative (CVIA 083-MAL99) through a grant to PATH (INV-007217) from the Bill and Melinda Gates Foundation. Research grants were made to the London School of Hygiene & Tropical Medicine, United Kingdom, with sub-contracts to the Malaria Research and Training Center (MRTC), Mali, the Institut de Recherche en Sciences de la Santé (IRSS) and to the University of Oxford.

## Data Availability Statement

Raw sequencing data generated in this study have been deposited in the NCBI Sequencing Read Archive with the accession PRJNA1345094.

## Author Contributions

DC and BG coordinated the clinical trial. AD, IS, AM, J-BO, IZ, and HT facilitated execution of the trial and collection of samples. KK generated the genotyping and validation data. EL analyzed the genotyping data and prepared all figures. DEN supervised genotyping data production and analysis. EL and DEN wrote the first draft of the paper, which BG reviewed and edited. All authors read and approved the final manuscript.

## Notes

### Competing Interest Statement

The authors have declared no competing interest.

### Author Declarations

We processed samples from the clinical trial NCT04319380, registered on ClinicalTrials.gov. The original trial protocol was reviewed and approved by the ethics committees of the London School of Hygiene & Tropical Medicine, UK; the University of Science, Techniques, and Technologies, Bamako, Mali; the ethics committee for health research, Burkina Faso; and the regulatory authorities in Burkina Faso and Mali.

### Summary of Updates

Author list corrected to include all contributing authors.

